# In-hospital mortality due to breakthrough COVID-19 among recipients of COVISHIELD (ChAdOx nCoV-19) and COVAXIN (BBV152)

**DOI:** 10.1101/2021.12.07.21267398

**Authors:** Tejas M Suri, Tamoghna Ghosh, M Arunachalam, Rohit Vadala, Saurabh Vig, Sushma Bhatnagar, Anant Mohan

## Abstract

**Background:** Multiple vaccines have received emergency-use authorization in different countries in the fight against the COVID-19 pandemic. India had started its vaccination campaign using the COVISHIELD (ChAdOx nCoV-19) and the COVAXIN (BBV152) vaccines. However, there is a lack of head-to-head comparisons of the different vaccines.

**Methods:** We performed a retrospective cohort study during the second wave of the pandemic in India with predominant circulation of the delta strain of SARS-CoV-2. We enrolled adult patients who were hospitalized with breakthrough COVID-19 infection after vaccination. We compared in-hospital outcomes between patients who had received the COVISHIELD (n=181) or COVAXIN vaccines.

**Results:** Between April and June 2021, a total of 353 patients were enrolled, among whom 181 (51.3%) received COVAXIN (156 partially vaccinated and 25 fully vaccinated) and 172 (48.7%) received COVISHIELD (155 partially vaccinated and 17 fully vaccinated). The in-hospital mortality did not differ between the recipients of COVISHIELD or COVAXIN in either the fully vaccinated [2 deaths (11.8%) vs 0 deaths (0%), respectively p=0.08] or the partially vaccinated cohorts [31 deaths (20%) vs 28 deaths (17.9%), respectively, p=0.65].

**Conclusions:** Patients who are hospitalized with breakthrough COVID-19 had similar in-hospital outcome irrespective of whether they received COVISHIELD or COVAXIN.

## Introduction

The COVID-19 pandemic has caused considerable morbidity and mortality with over 243 million cases and 4.9 million deaths worldwide as of October 2021.(1) The development of COVID-19 vaccines began in early 2020 and has progressed at breakneck speed.(2) Although several vaccines have received emergency use authorizations in different countries, their clinical trials have been of short durations. Furthermore, multiple vaccines have been simultaneously deployed by many countries. However, there is a lack of head-to-head comparisons of different vaccines.

The Indian vaccination programme began in January 2021 with the rollout of two vaccines, namely COVISHIELD (ChAdOx nCoV-19, an adenoviral vector vaccine) and COVAXIN (BBV152, a whole-virion inactivated vaccine). In an interim analysis of four randomized trials, the efficacy of two doses of ChAdOx nCoV-19 for preventing symptomatic COVID-19 was 70.4%.(3) The BBV152 was found to have an efficacy of 77.8% against symptomatic COVID-19.(4) These vaccines offer protection at a lower magnitude against the delta variant (B.1.617.2) of the causative severe acute respiratory syndrome coronavirus-2 (SARS-CoV-2), which was the predominant strain during the second wave of pandemic in India between April and June 2021.(5) The efficacy of ChAdOx nCoV-19 against the delta variant was found to be 67.0%, while that of BBV152 was 65.2%.(4,6) In this study conducted during the second wave of the pandemic in India, we compared the in-hospital outcomes of patients who were hospitalized with breakthrough infections of COVID-19 after vaccination with either COVISHIELD or COVAXIN.

## Methods

We retrospectively analysed a prospectively enrolled cohort of patients admitted to the COVID-19 facility at the National Cancer Institute, All India Institute of Medical Sciences, New Delhi. Patients aged above 18 years were included if they had breakthrough COVID-19 infection after vaccination with either COVISHIELD or COVAXIN. Patients were excluded if the final outcome was unknown due to transfer to another hospital or leave against medical advice. The patients were enquired regarding the type of vaccine received (COVAXIN vs COVISHIELD), the number of doses and the dates of vaccination. Patients were deemed fully vaccinated if they were hospitalized more than two weeks after receiving the second dose; whereas they were considered partially vaccinated if they had received either one dose or had received the second dose within two weeks prior to hospitalization. Baseline demographic characteristics and comorbidities were recorded. Disease severity at admission was classified into mild (no dyspnea or hypoxemia), moderate (presence of dyspnea, SpO_2_ between 90 and 94%, or respiratory rate between 24 and 30 breaths per minute) or severe COVID-19 (SpO2 less than 90% or respiratory rate greater than 30 breaths per minute).(7)

The in-hospital outcomes including mortality, need for invasive mechanical ventilation, need for high flow nasal oxygenation (HFNO)/non-invasive ventilation (NIV), and duration of hospital stay were compared between COVISHIELD and COVAXIN recipients in both the partially vaccinated and fully vaccinated subsets. Statistical analysis was performed using STATA v15. Comparisons between groups for continuous variables was performed using Student’s t-test, and that for categorical variables using chi-square test. The study protocol was approved by the hospital’s Institutional Ethics Committee.

## Results

Between April and June 2021, a total of 2055 patients were admitted, and outcome information was available for 1835 (Figure 1). The type of vaccine received was known for 353 patients, among whom 181 (51.3%) received COVAXIN (156 partially vaccinated and 25 fully vaccinated) and 172 (48.7%) received COVISHIELD (155 partially vaccinated and 17 fully vaccinated). Baseline characteristics of patients are presented in Table 1. COVISHIELD recipients were older than COVAXIN recipients [mean (SD) ages, 55.1 (14.8) vs 49.7 (17.0), p=0.01] and were more likely to have diabetes mellitus (31.6% vs 20.8%, p=0.02) and hypertension (32.2% vs 22.5%, p=0.04). COVISHIELD recipients were more likely than COVAXIN recipients to have moderate COVID-19 at presentation (29.0% vs 16.1%, p=0.01). There was no difference in the presence of severe disease between the two groups.

**Table 1.**
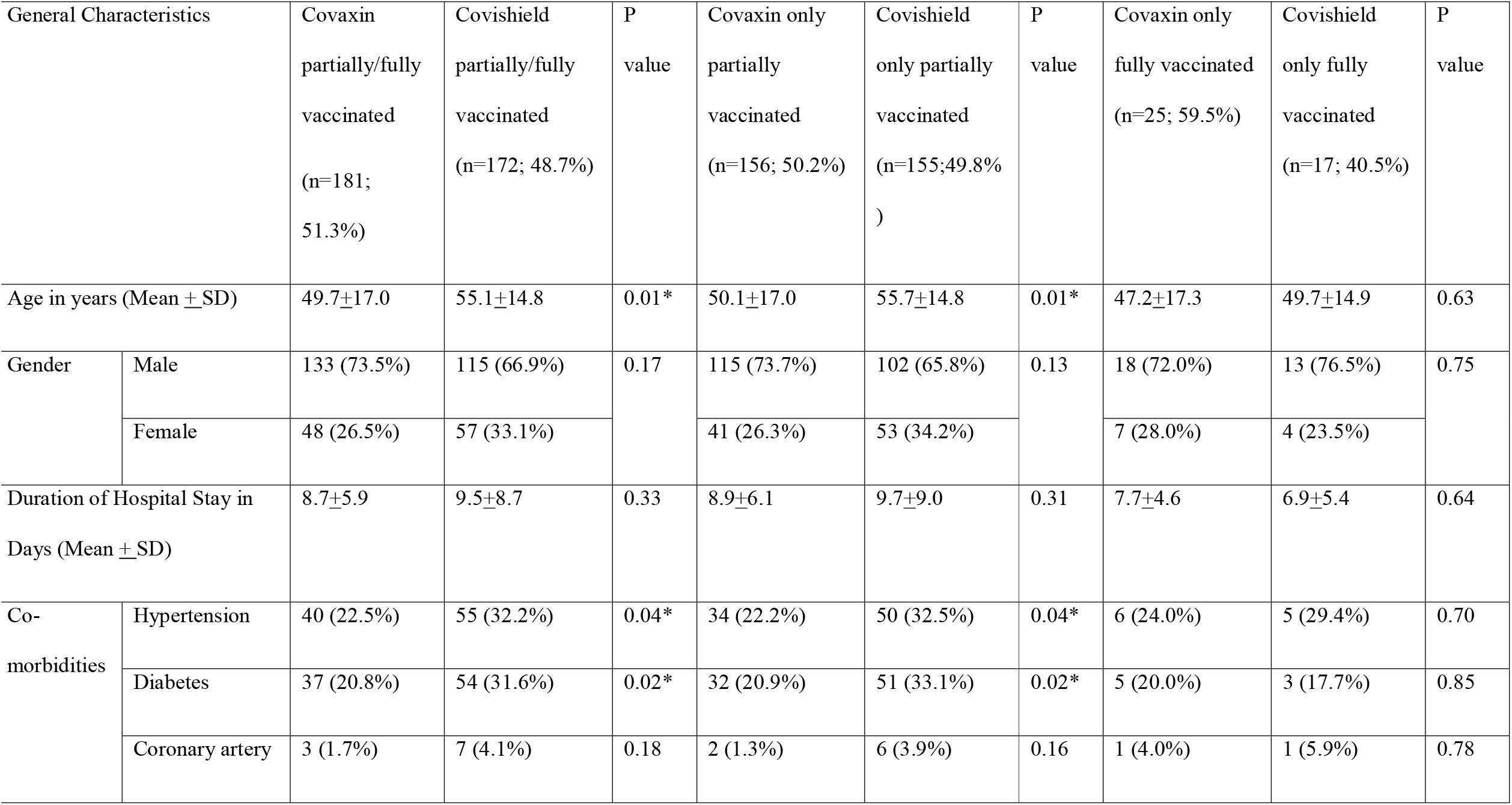
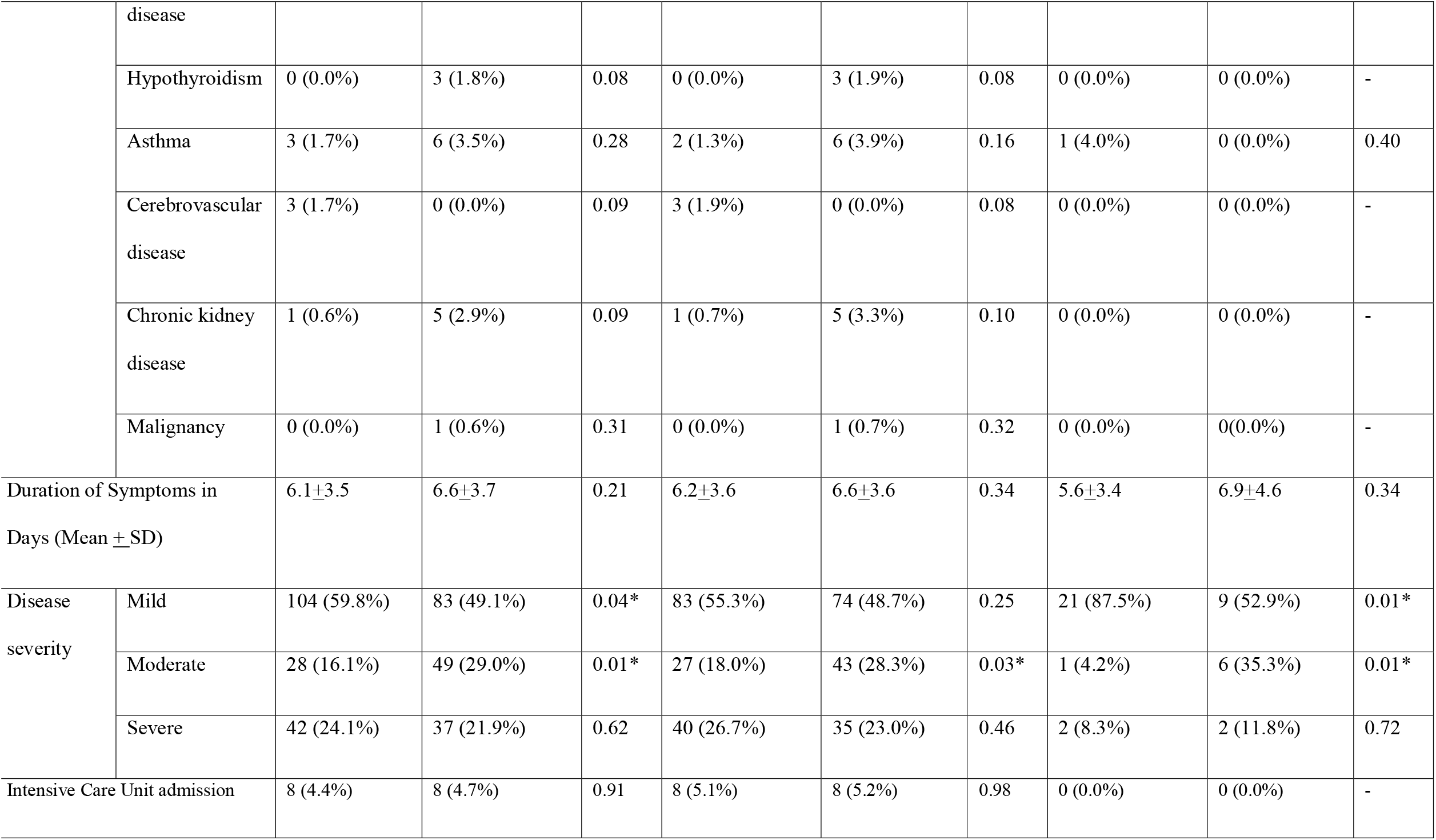
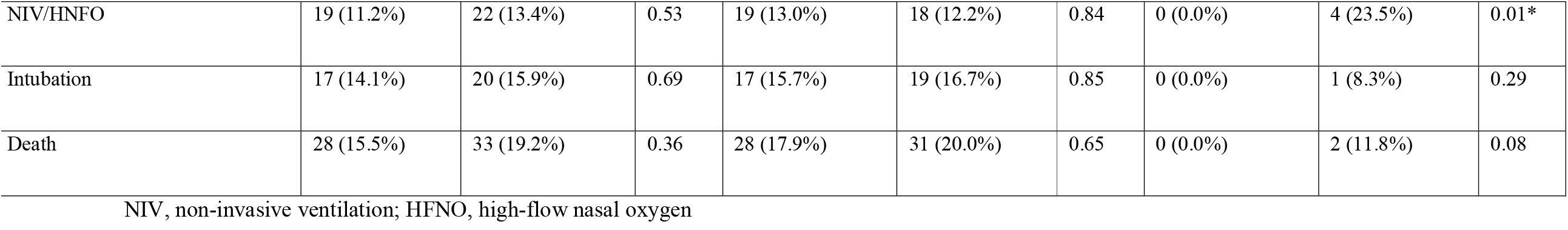
Baseline demographic and clinical characteristics of patients who received COVAXIN and COVISHIELD.

**Figure 1.**
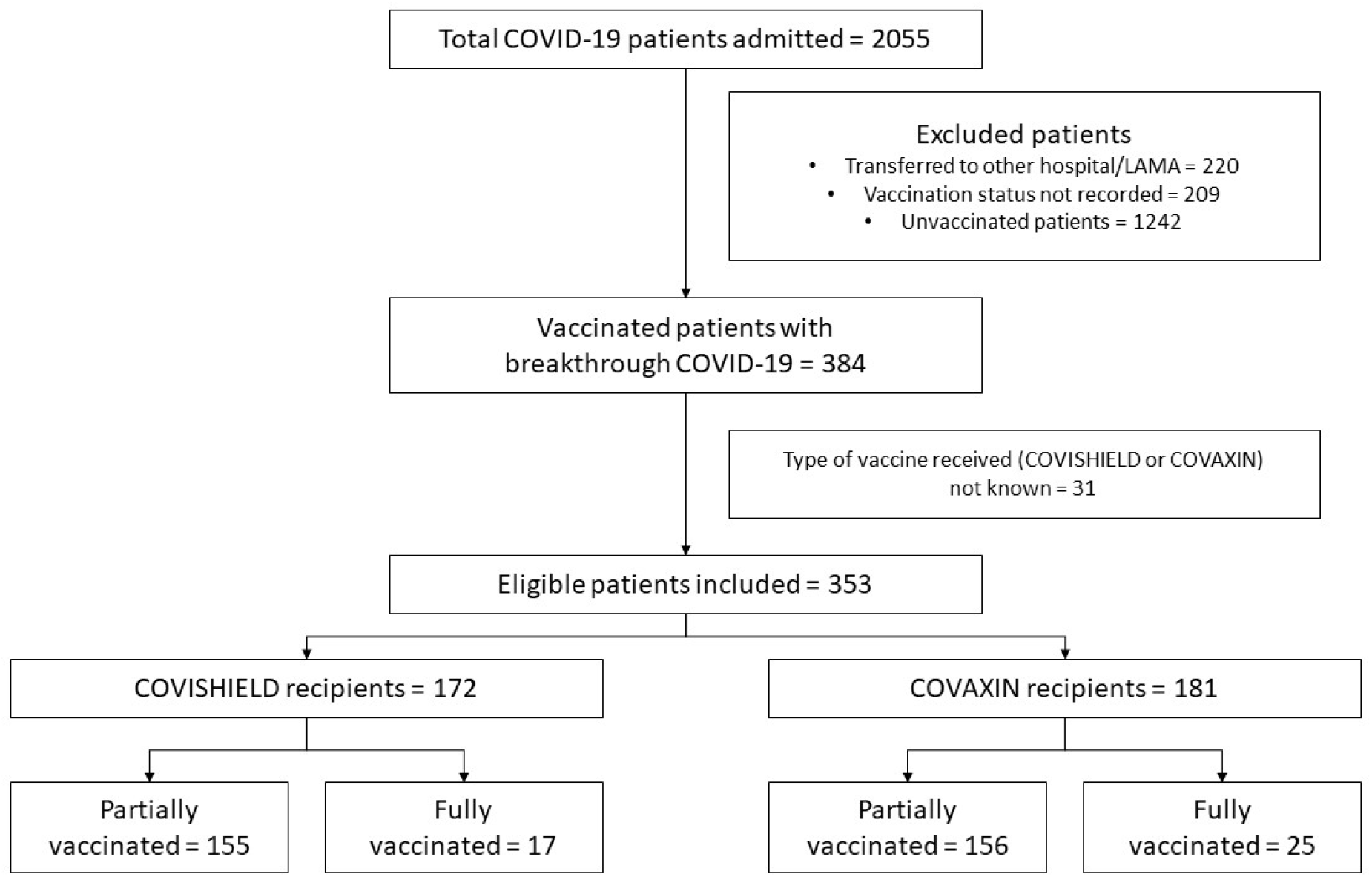
Study Flow.

The in-hospital mortality did not differ between the recipients of COVISHIELD or COVAXIN in either the fully vaccinated [2 deaths (11.8%) vs 0 deaths (0%), p=0.08] or the partially vaccinated cohorts [31 deaths (20%) vs 28 deaths (17.9%), p=0.65]. Furthermore, there was no difference in the need for mechanical ventilation or duration of hospital stay among recipients of either vaccine (Table 1). Fully vaccinated COVISHIELD recipients were more likely to require HFNO/NIV than COVAXIN recipients [4 patients (23.5%) vs 0 patients (0%), p=0.01].

## Discussion

### Main finding of this study

We found no difference in occurrence of severe disease at admission, need for mechanical ventilation or in-hospital mortality between patients with breakthrough COVID-19 who received either of the two vaccines (COVAXIN or COVISHIELD). Previously reported data from our hospital cohort demonstrated that mortality rates were significantly lower among fully vaccinated (5.7%) compared with partially vaccinated (19.5%) or unvaccinated (22.8%) individuals.(8) Taken together, these results are reassuring regarding the real-world efficiency of both vaccines in a setting with circulation of the delta variant of SARS-CoV-2.

### What is already known on this topic

In another study conducted among hospitalized COVID-19 patients in India, those who were fully vaccinated with COVISHIELD had a lower mortality than unvaccinated patients (12.5% vs 31.4%, p<0.0001).(9) Previously, Jain et al(10) had shown that the choice of vaccination (COVISHIELD vs COVAXIN) was considered to be important by medical students, with the former being preferred. This preference for COVISHIELD was influenced by a relative lack of efficacy data regarding COVAXIN at the time of initiation of India’s vaccination drive. However, subsequent reports demonstrating the efficacy of COVAXIN in preventing symptomatic COVID-19 should alleviate such concerns.(4,11)

### What this study adds

This is the first study to compare in-hospital outcomes of COVISHIELD and COVAXIN recipients who were hospitalized with breakthrough COVID-19 infection. We found no difference in the in-hospital mortality or need for mechanical ventilation between recipients of either vaccine.

### Limitations

The single-centre design and limited sample size are the major limitations of our study, which we consider as a preliminary report. Prospective studies with larger sample size are needed to establish the relative efficiencies of the available vaccines. Furthermore, the vaccination status was based on patient history which is subject to recall bias.

## Conclusion

In conclusion, our study suggests that both vaccines, i.e., COVISHIELD and COVAXIN, are equally likely to prevent death due to breakthrough COVID-19.

## Data Availability

Data will be made available on reasonable request

## Acknowledgements

The authors express their sincere gratitude to the data entry staff of Pulmonary, Critical Care and Sleep Medicine, All India Institute of Medical Sciences, New Delhi, India for their assistance in procuring the clinical and laboratory details and preparing the raw data.

